# Association between serum CEA levels and ctDNA-detected Epidermal Growth Factor Receptor mutations in lung adenocarcinoma

**DOI:** 10.64898/2026.07.14.26358115

**Authors:** Sharmistha Roy, Md Kabirul Islam Soroar, Hosne Ara, Sharif Al Nur, Riyadh Arifin Akanda, Subrata Saha, Mohammad Masum Alam

**Author notes:** **Corresponding Author:** Sharmistha Roy.

## Abstract

**Background with objective:** Detecting EGFR mutations is critical for treating lung adenocarcinoma with highly effective targeted therapies. However, standard genetic testing is expensive, complex, and often unavailable in resource-limited settings like Bangladesh. Because elevated serum CEA has been linked to these genetic alterations, it could serve as an accessible screening tool. This study aims to evaluate the association between serum CEA levels and EGFR mutation status to determine if routine CEA testing can reliably predict these mutations and guide treatment.

**Methodology:** In this cross-sectional analytical study, we recruited 58 patients with histologically confirmed treatment naïve lung adenocarcinoma. The presence of EGFR mutations in the ctDNA was determined via ARMS (Amplification Refractory Mutation System) PCR. Patient data was statistically analyzed to assess the diagnostic correlation between serum CEA levels and the presence of EGFR mutations.

**Result:** The overall EGFR mutation rate was 43.1% with exon 19 deletion (48%) and exon 21 mutations (44%) were the predominant types. Median serum CEA levels were significantly higher in patients with EGFR mutations compared to wild-type cases (14.6 ng/ml vs 2.8 ng/ml, p<0.001). A multivariate analysis revealed a 14% increased likelihood of an EGFR mutation for 1 ng/ml rise in serum CEA. Furthermore, serum CEA showed strong diagnostic accuracy for ctDNA samples at a 6.39 ng/ml cut-off (AUC 0.82, sensitivity 68.0%, specificity 84.8%).

**Conclusion:** Serum CEA is a valuable, cost-effective, and non-invasive biomarker demonstrating significantly higher levels and strong diagnostic accuracy in EGFR-mutated lung adenocarcinoma compared to wild-type cases.

## INTRODUCTION

Malignancies develop through abnormal changes in the natural process of cell division ^1^. In 2022, lung malignancies remained the predominant cause of cancer-driven mortalities worldwide, accounting for 1.8 million deaths.^2^ In Bangladesh, most lung cancer cases are diagnosed at an advanced stage, contributing to poor survival rates.^3^ Approximately 85% of all lung malignancies are classified as non-small cell lung cancer (NSCLC), with adenocarcinoma being the primary subtype.^4^

Lung adenocarcinoma is frequently driven by genetic alterations in genes such as EGFR, KRAS, and ALK.^5^ Among these, EGFR mutations are notably highly among Asian adenocarcinoma patients, presenting in as many as 51.4% of case.^6^ Identifying these mutations is critical because they reliably predict tumor sensitivity to EGFR tyrosine kinase inhibitors (EGFR-TKIs).^7^ TKIs have revolutionized the lung cancer management with better progression free survival and significantly lower toxicity than traditional chemotherapy.^8,9^

EGFR is a single-pass transmembrane protein made of 1210 amino acids. It has five regions: an outer extracellular region, a lipid-soluble transmembrane section, a juxtamembrane domain, an interior kinase component, and a regulatory tail at the C-terminal.^10,11^ Activation occurs when a naturally occurring ligand, like the epidermal growth factor (EGF), binds to the extracellular domain. It causes the C-terminal tail of the receptor to become tyrosine-phosphorylated. This phosphorylation subsequently activates several downstream signaling mechanisms. These include the RAS/RAF/MAPK pathway, PI3K/Akt/mTOR pathway, and JAK/ STAT3 pathway.^12,13^ Mutations are most frequently identified within the tyrosine kinase domain which consists of exons 18, 19, 20, and 21. Mutations that activate the receptor or disruptions in any of these mechanisms can lead to the continuous activation of EGFR, even without ligand binding. This constant activation can promote cell proliferation, survival, and spread, contributing to cancer development.^14^

While identifying EGFR mutations is now a clinical mandate for treating advanced lung adenocarcinoma, widespread genetic testing remains a significant hurdle. The molecular profiling and next-generation sequencing required for these diagnoses are complex and expensive. In resource-limited settings like Bangladesh, these advanced diagnostics are simply unaffordable for most patients and are rarely available outside a few specialized centers.^15,16^ Consequently, there is an urgent need for a simple, cost-effective screening tool to help clinicians identify which patients are most likely to harbor EGFR mutations, thereby ensuring that the high expense of genetic testing is directed toward those who will truly benefit.^8,9^

Serum tumor markers offer a highly accessible and reliable solution to this problem.^17^ Carcinoembryonic antigen (CEA), a cell surface glycoprotein involved in abnormal cell adhesion and metastasis, is already widely utilized in routine lung cancer management.^9^ Clinical evidence shows a strong link between elevated serum CEA levels and positive treatment responses to EGFR-targeted therapies. This suggests that routine CEA testing could serve as an effective pre-screening marker for EGFR mutations. Validating this correlation would establish a practical diagnostic pathway, improving personalized treatment plans while addressing the socioeconomic challenges patients face in Bangladesh.

When it does come time to confirm these genetic mutations, traditional protocols dictate using tissue biopsies containing at least 20% malignant cells.^18^ However, this study exclusively utilized circulating tumor DNA (ctDNA) instead of tissue biopsy for several practical and biological reasons. Most lung adenocarcinoma patients present at advanced stages when the patient is too frail for invasive surgical sampling.^19^ Besides, invasive biopsies carry severe complication risks, such as pneumothorax and hemoptysis,^20^ and up to 30% fail to yield enough tissue for molecular analysis.^18^ Furthermore, a localized tissue sample can miss the overall genetic heterogeneity of the cancer. By utilizing ctDNA, a clinically approved, non-invasive liquid biopsy,^21^ we safely captured a much broader genetic landscape of the tumor burden. Crucially, because both serum CEA and ctDNA are evaluated from peripheral blood, it eliminates the need to compare a blood marker against a tissue biopsy. By establishing this non-invasive, blood-to-blood correlation, we hope to validate serum CEA as an accessible first-line screening tool that can confidently predict the need for advanced ctDNA analysis.

## MATERIALS AND METHODS

### Study design and participants

This analytical, cross-sectional research was conducted at the Department of Biochemistry and Molecular Biology, Bangabandhu Sheikh Mujib Medical University (currently known as Bangladesh Medical University), Dhaka, spanning from March 2024 to February 2025. Ethical permission was granted from the university’s Institutional Review Board (IRB). Using a non-probability sampling technique, 58 histologically confirmed lung adenocarcinoma patients were selected from the oncology departments of several tertiary care hospitals. Inclusion criteria required patients to be at least 40 years old with a histologically confirmed diagnosis of primary lung adenocarcinoma. Exclusion criteria included prior anti-cancer therapy or surgery, secondary metastatic lesions from other organs, pregnant or lactating women, as well as patients with a history of liver cirrhosis, inflammatory bowel disease, or pancreatitis. Following the acquisition of informed written consent, patients underwent an initial clinical evaluation through detailed history-taking. These baseline clinical details, along with all subsequent laboratory reports, were then documented using a standardized data collection form.

### Mutation Analysis and CEA testing

To isolate ctDNA, 8 mL samples of venous blood were collected in K2EDTA vacutainers and processed within a two-hour window. Plasma was separated using a dual-centrifugation protocol (3,000 × g followed by 11,000 × g, each for 10 minutes) and stored at -80 °C. We extracted cfDNA using the CatchGene® Catch-cfDNA Serum/Plasma 1000 Kit, confirming DNA purity (target A260/A280 ratio: 1.7–2.0) with a NanoDrop™ ND 2000 Spectrophotometer (Thermo Fisher Scientific Inc). To detect EGFR mutations, we used the AmoyDx EGFR 29 Mutations Detection Kit (Amoy Diagnostics, Xiamen, China) paired with a Rotor-Gene Q PCR cycler (Qiagen, Hilden, Germany). Based on Amplification Refractory Mutation System (ARMS) technology, this assay employs specific fluorescent probes to qualitatively identify 29 somatic mutations across EGFR exons 18 to 21 (including G719X, E19 deletions, T790M, S768I, exon 20 insertions, L858R, and L861Q). The 46-cycle PCR protocol incorporated both internal and external controls to verify template quality and rule out PCR inhibitors. Mutation status was determined by calculating the ΔCt value (Mutant FAM Ct minus External Control FAM Ct). Samples were classified as mutation-positive if the mutant FAM Ct was < 29 and ΔCt < 13. Any sample with a ΔCt ≥ 13 or lacking amplification was reported as negative. Serum CEA levels were estimated using a two-site sandwich immunoassay with direct chemiluminometric technology on the Atellica Solution IM Analyzer (Siemens Healthineers, Germany).

### Statistical Analysis

Data were analyzed using IBM SPSS Statistics for Windows, version 27.0 (IBM Corp., Armonk, N.Y., USA). Continuous variables were presented as mean (±SD) or median (IQR) and compared using the unpaired Student’s t-test or Mann-Whitney U test, based on distribution. Categorical variables were reported as frequencies (%) and analyzed via the chi-square test. We used multivariate logistic regression to assess the association between serum CEA and EGFR mutations, and ROC curve analysis (AUC with 95% CI) to evaluate CEA’s diagnostic performance. Supplementary statistical computing and data visualization were conducted using Python (version 3.10) with the pandas, scikit-learn, and matplotlib libraries. Statistical significance was defined as p < 0.05.

## RESULTS

### Baseline characteristics of the study population

This study included 58 patients with lung adenocarcinoma. Key baseline demographic and clinical parameters are outlined in Table 1. Participants had an average age of 59.8 (± 10.4) years. Most patients were male (n = 42, 72.4%) and ex-smokers (n = 31, 53.4%). The majority presented with Stage IV disease (n = 34, 58.6%) and moderately differentiated tumors (n = 37, 63.8%). EGFR mutations were detected by ctDNA in 25 participants (43.1%), while 33 (56.9%) had wild-type EGFR. Among the 25 ctDNA-positive cases, exon 19 deletions (n = 12) and L858R substitutions (n = 11) were the most commonly observed mutations, alongside single occurrences of T790M (n = 1) and S768I (n = 1) mutations.

**Table 1:** Baseline Characteristics and Clinical Demographics Categorized by EGFR Mutation (N=58)

| Characteristics | EGFR Mutated<br>($n$ = 25) | EGFR Wild<br>($n$ = 33) | Total<br>($N$ = 58) |
| --- | --- | --- | --- |
| **Age** |  |  |  |
| Mean ± *SD* | 59.2 ± 11.7 | 60.2 ± 9.4 | 59.8 ± 10.4 |
| ≤ 60 years, $n$ (%) | 11 (44.0) | 18 (55.6) | 29 (50.0) |
| $>$ 60 years, $n$ (%) | 14 (56.0) | 15 (45.4) | 29 (50.0) |
| **Gender** |  |  |  |
| Male, $n$ (%) | 16 (64.0) | 26 (78.8) | 42 (72.4) |
| Female, $n$ (%) | 9 (36.0) | 7 (21.2) | 16 (27.6) |
| **Smoking status** |  |  |  |
| Ex-smoker, $n$ (%) | 11 (44.0) | 20 (60.6) | 31 (53.4) |
| Non-smoker, $n$ (%) | 14 (56.0) | 13 (39.4) | 27 (46.6) |
| **Disease stage** |  |  |  |
| Stage II-III, $n$ (%) | 4 (16.0) | 20 (60.6) | 24 (41.4) |
| Stage IV, $n$ (%) | 21 (84.0) | 13 (39.4) | 34 (58.6) |
| **Histological grading** |  |  |  |
| Moderately differentiated, $n$ (%) | 20 (80.0) | 17 (51.5) | 37 (63.8) |
| Poorly differentiated, $n$ (%) | 5 (20.0) | 16 (48.5) | 21 (36.2) |

*Note.* EGFR = epidermal growth factor receptor; *SD* = standard deviation. Mutation status was detected by ctDNA.

### Evaluating serum CEA concentrations in relation to EGFR mutations

Serum CEA levels were compared based on EGFR mutation status (Table 2). The median serum CEA level was significantly elevated in patients with EGFR-mutated lung adenocarcinoma (14.6 ng/ml; IQR: 5.1–21.4) compared to those with wild-type EGFR (2.8 ng/ml; IQR: 1.4–4.1) (*p* < .001).

**Table 2:** Comparison of serum CEA levels between EGFR-mutated and wild-type lung adenocarcinoma.

| EGFR Mutation Status | n (%) | Serum CEA Median (IQR) | p |
| --- | --- | --- | --- |
| Mutated | 25 (43.1) | 14.6 (5.1, 21.4) | < .001 |
| Wild | 33 (56.9) | 2.8 (1.4, 4.1) |  |

*Note.* CEA = carcinoembryonic antigen; IQR = interquartile range. The $p$ -value was calculated using the Mann-Whitney U test.

### Correlation of CEA with EGFR mutation

We performed a multivariate logistic regression to isolate independent variables correlated with the presence of EGFR mutations (Table 3). After adjusting for age, gender, and smoking status, serum CEA stood out as the sole independent predictive factor. For every 1 ng/ml increase in serum CEA, there was a corresponding 14% increase in the probability of detecting an EGFR mutation (OR = 1.14; 95% CI [1.04, 1.26]; p = .004).

**Table 3:** Multivariate logistic regression analysis of factors associated with EGFR mutation.

| Variable | aOR | 95% CI | p |
| --- | --- | --- | --- |
| Serum CEA level | 1.14 | [1.04, 1.26] | .004 |
| Age | 1.00 | [0.96, 1.06] | .963 |
| Gender (Female) | 4.78 | [0.62, 36.90] | .134 |
| Smoking status (Never) | 0.42 | [0.06, 2.81] | .374 |

*Note.* aOR = adjusted odds ratio; CI = confidence interval; CEA = carcinoembryonic antigen. Multivariate logistic regression analysis was performed to adjust for covariates. All precision estimates are presented with a 95% confidence interval.

### Diagnostic performance and ROC curve

The diagnostic performance of serum CEA for predicting EGFR mutations is illustrated in Figure 1. Serum CEA demonstrated strong diagnostic accuracy, yielding an area under the curve (AUC) of 0.82 (95% CI [0.71, 0.92]). At the optimal triage cut-off value of 6.39 ng/ml, diagnostic evaluation revealed a sensitivity of 68.0%, specificity of 84.8%, positive predictive value (PPV) of 77.3%, and negative predictive value (NPV) of 77.8%.

**Figure 1:**
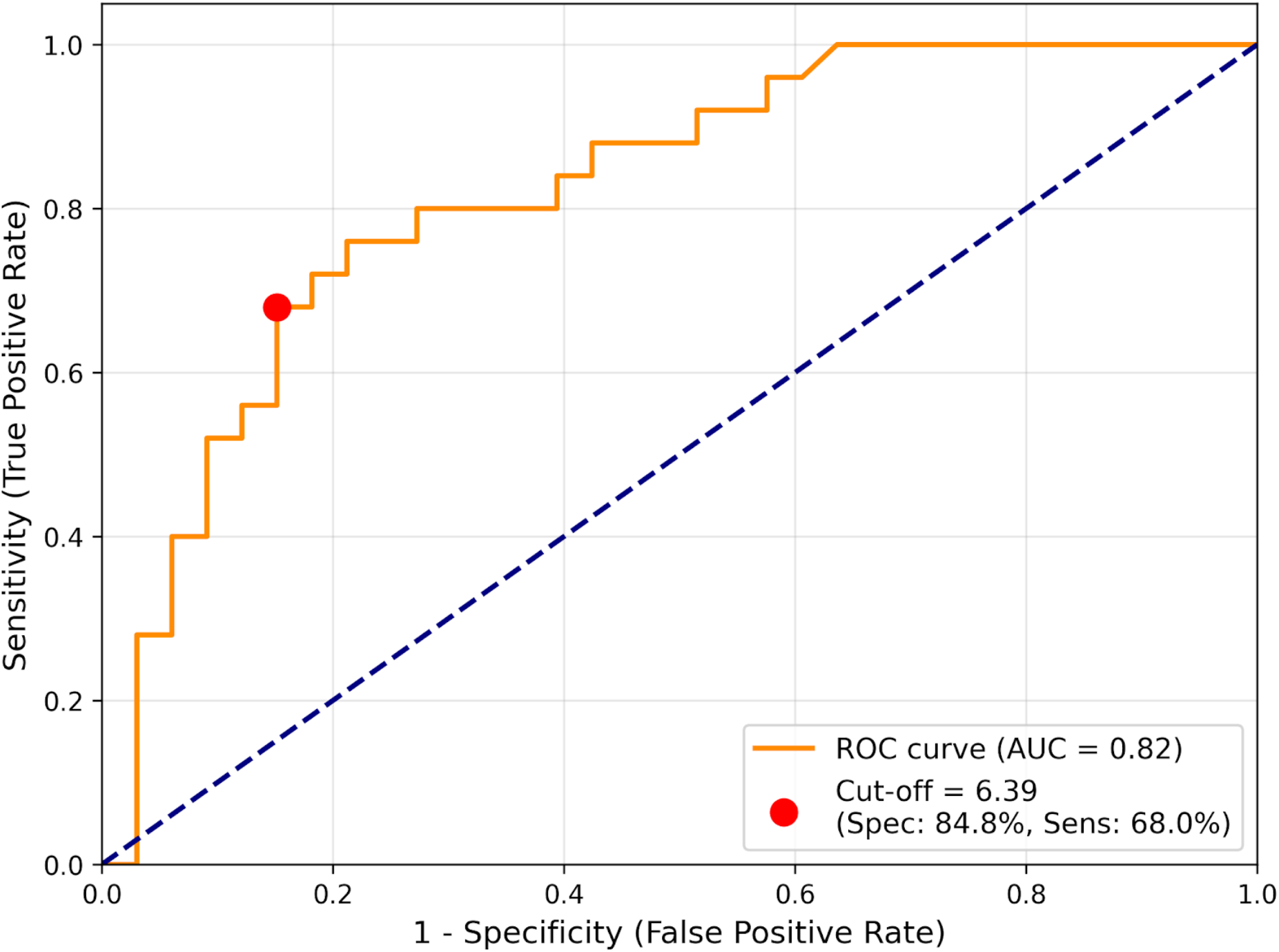
Receiver Operating Characteristic (ROC) Curve of Serum CEA for the Prediction of EGFR Mutation.

## DISCUSSION

Obtaining conventional tissue biopsies is frequently impractical in advanced NSCLC. Therefore, non-invasive ctDNA analysis provides a reliable diagnostic alternative. To the best of our knowledge, this is the first Bangladeshi study to evaluate serum CEA as a predictive indicator for EGFR mutations detected through ctDNA in lung adenocarcinoma. Our study identified a overall EGFR mutation rate of 43.10% and demonstrated that elevated serum CEA levels act as a significant, independent predictor of these mutations, exhibiting a clear dose-dependent relationship and high diagnostic specificity.

In our study, the 43.10% mutation rate represents a notable increase from the 23% prevalence reported in Bangladesh a decade ago.^22^ This increase likely reflects the enhanced sensitivity of modern ARMS-PRC methods, alongside potential shifts in environmental exposures. Consistent with global literature, exon 19 deletions and exon 21 L858R point mutations accounted for 92% of all alterations.^23,24^ Demographically, mutations were classically more prevalent in females and non-smokers.^8^ However, our multivariate analysis did not identify gender or smoking status as independent predictors. This aligns with adjusted analyses by Kosaka et al.,^25^ who similarly found that gender and smoking lost independent predictive value when adjusted for other variables.

In this study we found that, patients with ctDNA-detected EGFR mutations exhibited substantially higher median serum CEA levels (14.6 ng/mL) compared to wild-type patients (2.8 ng/mL). Multivariate analysis confirmed a 14% increased likelihood of an EGFR mutation for each unit increase in CEA (*p* = .004). This supports previous global findings that elevated CEA acts as a reliable prognostic indicator for these targetable mutations.^23,26–28^ Beyond statistical correlation, this relationship is rooted in tumor biology. The constitutive activation of the mutated EGFR signaling pathway drives aggressive tumor proliferation and metabolic activity, which is clinically mirrored by the overexpression and systemic release of CEA in lung adenocarcinoma cells.^29^ Our liquid biopsy findings validate this biological mechanism, demonstrating that circulating CEA accurately reflects the underlying genetic mutation burden.

Furthermore, we observed a clear trend of increasing EGFR mutation rates with rising serum CEA levels. The mutation rate was 18.75% in patients with normal CEA (< 5 ng/mL), rising to 33.33% for levels between 5–20 ng/mL, and peaking at 87.50% in patients with CEA > 20 ng/mL. This perfectly mirrors the findings reported by Yang et al.^8^ They reported EGFR mutation rates of 18.75%, 36.36%, and 62.50% for serum CEA levels of <5 ng/mL, 5–19 ng/mL, and ≥20 ng/mL, respectively, with a peak mutation rate of 85.71% at CEA levels of 20–49 ng/mL. Conversely, the predictive utility of serum CEA may be highly dependent on regional and demographic factors. A recent Caucasian cohort study found no significant association between CEA and EGFR mutation status.^16^ This highlights a major difference between Western and Asian populations. Western groups have naturally lower EGFR mutation rates (10–15%) and much higher smoking rates. Because smoking raises CEA levels on its own, it can easily hide the CEA increases caused by genetic mutations.^16^

Beyond general correlation, our receiver operating characteristic (ROC) analysis established the direct clinical utility of this biomarker. At an optimal cut-off value of 6.39 ng/mL, representing a threshold slightly above the standard clinical upper limit of normal (5.0 ng/mL) used in routine laboratory settings, serum CEA yielded a strong area under the curve (AUC) of 0.82. While our sensitivity (68.0%) was slightly lower than cohorts evaluated in China (69.6%) and Vietnam (76.5%), our specificity of 84.8% vastly outperformed the 48.8% and 47.5% reported in those respective studies. ^23,30^ This exceptional specificity is particularly noteworthy when placed in a broader context. For instance, a comprehensive meta-analysis of Asian cohorts by Zhang et al.^31^ reported a pooled specificity for CEA of only 66%. Our study’s elevated specificity indicates a substantially lower rate of false-positive predictions in the Bangladeshi demographic compared to other regional populations.

## CONCLUSION

Elevated serum CEA levels are significantly and independently associated with ctDNA-detected EGFR mutations in lung adenocarcinoma. Implementing routine serum CEA testing offers a highly specific, non-invasive, and cost-effective preliminary screening strategy. By identifying patients with the highest probability of harboring targetable mutations, clinicians can optimize the deployment of advanced liquid biopsy testing, ultimately accelerating access to personalized targeted therapies in resource-constrained settings.

## LIMITATIONS

This study has few limitations. Our cross-sectional design and small sample size from local tertiary hospitals limit how widely these findings can be generalized. Because our cohort reflects a specific demographic in Bangladesh, the optimal CEA cut-offs we identified may differ for populations with other genetic or environmental backgrounds. Additionally, without a longitudinal follow-up, we could not assess how elevated CEA impacts overall survival or patient response to EGFR-TKI therapy. Finally, as CEA is not entirely cancer-specific, unrecorded benign inflammation or lifestyle factors may have influenced some measurements.

## RECOMMENDATIONS

Given its high specificity and low cost, routine serum CEA testing should be integrated into clinical evaluations for advanced lung adenocarcinoma. Future research should also explore combining CEA with other biomarkers (such as CYFRA 21-1) to boost sensitivity, and use longitudinal tracking to determine if baseline CEA levels can accurately predict long-term EGFR-TKI treatment outcomes.

## Author Contributions

**Sharmistha Roy:** Conceptualization, Data curation, Formal analysis, Investigation, Methodology, Resources, Writing - original draft, Writing - review & editing. **Md Kabirul Islam Soroar:** Data curation, Formal analysis, Investigation, Methodology, Resources, Software, Writing - original draft, Writing - review & editing. **Hosne Ara:** Data curation, Formal analysis, Investigation, Writing - review & editing. **Sharif Al Nur:** Data curation, Formal analysis, Writing - review & editing. **Riyadh Arifin Akanda:** Data curation, Formal analysis, Writing - review & editing. **Subrata Saha:** Data curation, Formal analysis, Writing - review & editing. **Mohammad Masum Alam:** Conceptualization, Formal analysis, Supervision, Writing - review & editing. All authors reviewed and endorsed the final version of the manuscript and accept responsibility for the integrity and accuracy of the work.

## STATEMENTS AND DECLARATIONS

### Ethical approval

This study received ethical clearance from the Institutional Review Board of Bangabandhu Sheikh Mujib Medical University (Registration No. 5113, approval No: BSMMU/2024/6769, approval date: 13.07.2024). All research activities adhered to the principles of the Declaration of Helsinki and applicable national regulations. Participant information was anonymized before analysis to safeguard confidentiality.

### Consent to participate

Written informed consent was obtained from each participant using a standardized form, following a thorough explanation of the study objectives and procedures.

### Declaration of conflicting interest

The authors declare no competing interests related to this research, authorship, or publication.

### Funding statement

No external funding was received for the conduct of this study or preparation of the manuscript.

### Data availability

The datasets generated and/or analyzed during the study can be accessed from the corresponding author upon reasonable request.

## Notes

### Competing Interest Statement

The authors have declared no competing interest.

### Author Declarations

The Institutional Review Board of Bangabandhu Sheikh Mujib Medical University gave ethical approval for this work (Registration No: 5113, approval No: BSMMU/2024/6769, approval date: 13.07.2024).

